# Proteomic Characterization of Acute Kidney Injury in Patients Hospitalized with SARS-CoV2 Infection

**DOI:** 10.1101/2021.12.09.21267548

**Authors:** Ishan Paranjpe, Pushkala Jayaraman, Chen-Yang Su, Sirui Zhou, Steven Chen, Ryan Thompson, Diane Marie Del Valle, Ephraim Kenigsberg, Shan Zhao, Suraj Jaladanki, Kumardeep Chaudhary, Steven Ascolillo, Akhil Vaid, Arvind Kumar, Edgar Kozlova, Manish Paranjpe, Ross O’Hagan, Samir Kamat, Faris F. Gulamali, Justin Kauffman, Hui Xie, Joceyln Harris, Manishkumar Patel, Kimberly Argueta, Craig Batchelor, Kai Nie, Sergio Dellepiane, Leisha Scott, Matthew A Levin, John Cijiang He, Mayte Suarez-Farinas, Steven G Coca, Lili Chan, Evren U Azeloglu, Eric Schadt, Noam Beckmann, Sacha Gnjatic, Miram Merad, Seunghee Kim-Schulze, Brent Richards, Benjamin S Glicksberg, Alexander W Charney, Girish N Nadkarni

## Abstract

Acute kidney injury (AKI) is a known complication of COVID-19 and is associated with an increased risk of in-hospital mortality. Unbiased proteomics using biological specimens can lead to improved risk stratification and discover pathophysiological mechanisms. Using measurements of ∼4000 plasma proteins in two cohorts of patients hospitalized with COVID-19, we discovered and validated markers of COVID-associated AKI (stage 2 or 3) and long-term kidney dysfunction. In the discovery cohort (N= 437), we identified 413 higher plasma abundances of protein targets and 40 lower plasma abundances of protein targets associated with COVID-AKI (adjusted p <0.05). Of these, 62 proteins were validated in an external cohort (p <0.05, N =261). We demonstrate that COVID-AKI is associated with increased markers of tubular injury (NGAL) and myocardial injury. Using estimated glomerular filtration (eGFR) measurements taken after discharge, we also find that 25 of the 62 AKI-associated proteins are significantly associated with decreased post-discharge eGFR (adjusted p <0.05). Proteins most strongly associated with decreased post-discharge eGFR included desmocollin-2, trefoil factor 3, transmembrane emp24 domain-containing protein 10, and cystatin-C indicating tubular dysfunction and injury. Using clinical and proteomic data, our results suggest that while both acute and long-term COVID-associated kidney dysfunction are associated with markers of tubular dysfunction, AKI is driven by a largely multifactorial process involving hemodynamic instability and myocardial damage.

## Introduction

Severe acute respiratory syndrome coronavirus 2 (SARS-CoV-2) is a novel coronavirus that has caused the coronavirus disease 2019 (COVID-19) pandemic. Although effective vaccines are available, novel variants that may evade neutralizing antibodies exist in the population and have led to high case counts and periodic case surges. COVID-19 most commonly presents with fever, cough, and dyspnea^1, 2^ and is associated with acute respiratory distress syndrome (ARDS). However, the clinical syndrome resulting from SARS-CoV-2 infection is broad, ranging from asymptomatic infection to severe disease with extrapulmonary manifestations^3^, including acute kidney injury^4^, acute myocardial injury^5, 6^ and thrombotic complications^7–11^. The CRIT-COV-U research group in Germany recently developed a urinary proteomics panel COV50 that could consider this variability in infection by generating biomarkers that can indicate adverse COVID-19 outcomes based on the WHO severity scale^12^

Acute kidney injury (AKI) is a particularly prominent complication. The rates of AKI vary greatly based on patient population, but evidence suggests that at least 30% of hospitalized patients and 50% of patients in the intensive care unit (ICU) develop AKI^1, 4, 13–16^. Although the rate of AKI in hospitalized COVID-19 patients has decreased since the initial surge in 2020, the incidence remains high^17^. Like community-acquired pneumonia^18^, AKI is increasingly recognized as a common complication of COVID-19 in the hospitalized setting and confers significantly increased morbidity and mortality^19^.

There is a limited understanding of the pathophysiology of COVID-19-associated AKI. A recent paper^20^ compared transcriptomics and proteomics of postmortem kidney samples of patients with severe COVID-19 and autopsy-derived control cohorts of sepsis-AKI and non-sepsis-AKI. The work found common inflammatory pathways and regulatory responses including the downregulation of oxidative signaling pathways between COVID-19 AKI and sepsis-AKI. They also confirmed the observation of tubular injury in almost all their COVID-19 AKI samples while drawing similarities between the inflammation response of sepsis-associated AKI and COVID-19 associated AKI. Histopathological reports from autopsy specimens have provided conflicting insights into the pathological changes in the kidney in COVID-19. A report of 26 patients who died with COVID-19 AKI revealed acute tubular injury as a prominent mechanism^21^. Additionally, the presence of viral particles in the tubular epithelium and podocytes in autopsy specimens has been reported^21, 22^, which is evidence of direct viral invasion of the kidney. In addition, coagulopathy and endothelial dysfunction are hallmarks of COVID-19^23^ and may also contribute to AKI. Finally, SARS-CoV-2 may directly activate the complement system^24^. In addition to these mechanisms, systemic effects of critical illness (hypovolemia, mechanical ventilation) and derangements in cardiac function and volume may also contribute to COVID-19 AKI.

In addition to morbidity and mortality in the acute setting, COVID-19 is also associated with long term manifestations i.e., the post-acute sequelae of SARS-CoV2 (PASC)^25^. Kidney function decline is a major component of PASC and a study of more than 1 million individuals found that survivors of COVID-19 had an elevated risk of post-acute eGFR decline^26^, suggesting long term kidney dysfunction may occur following the acute infection.

Given the high incidence of COVID-19 associated kidney dysfunction, the unknown pathophysiology, and the urgent need for better approaches for risk stratification for long term kidney function decline we aimed to characterize the proteomic changes associated with COVID associated AKI and long-term kidney function. Proteomic biomarkers have previously shown success in predicting COVID-19 outcomes^27–29^. Other work^12^ has also applied urinary proteomic profiling to predict worsening of COVID-19 at early stages of the infection. Prior research using minimally invasive proteomics assays supports the use of peripheral serum as a readily accessible source of proteins that accurately reflect the human disease state^30–33^. We measured protein expression of more than 4000 proteins from serum samples collected in a diverse large cohort of hospitalized patients with COVID-19 and validated significant results in an independent cohort and identified proteins that are significantly different between patients with and without AKI. We then determined whether these proteomic perturbations also characterize post-discharge kidney function decline as measured by estimated glomerular filtration rate (eGFR).

## MATERIALS AND METHODS

### Patient cohort

An overview of the discovery cohort selection process is provided in **Fig 1**. We prospectively enrolled patients hospitalized with COVID-19 between March 24, and August 26, 2020, at five hospitals of a large urban, academic hospital system in New York City, NY into a cohort as previously described^34^. The cohort enrolled patients who were admitted to the health care system with a COVID-19 infection and had broad inclusion criteria without specific exclusion criteria. The Mount Sinai Institutional Review Board approved this study under a regulatory approval allowing for access to patient level data and biospecimen collection^35^. Peripheral blood specimens were collected at various points during the hospital admission for each patient.

**Fig 1:**
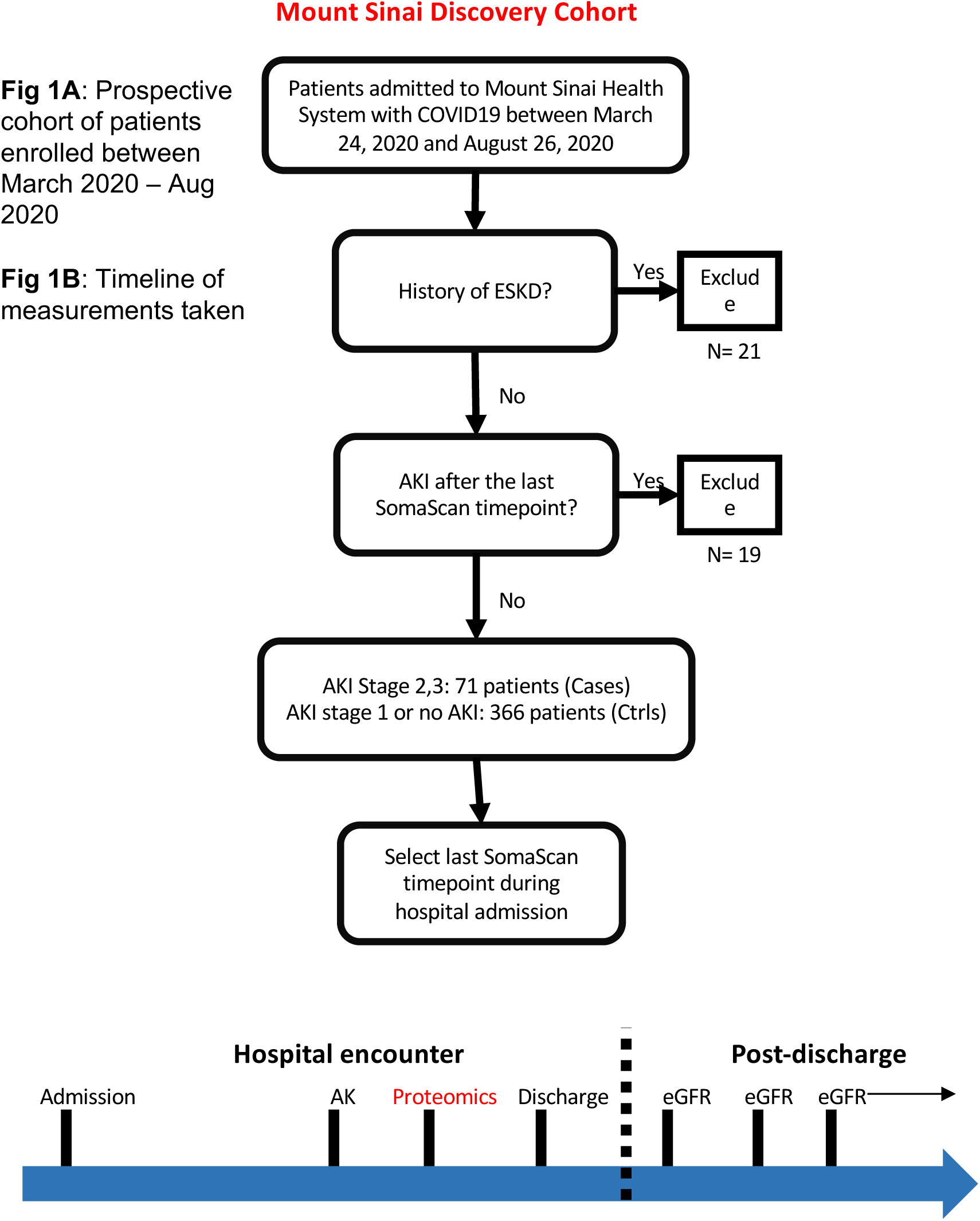
Overview of the discovery cohort selection process.

The validation cohort included a prospective biobank from Quebec, Canada that enrolled patients hospitalized with COVID-19, as previously described^29^. Patients were recruited from the Jewish General Hospital and Centre Hospitalier de l’Université de Montréal. Peripheral blood specimens were collected at multiple time points after admission.

We defined an AKI cohort using proteomic data acquired at the last available timepoint during the hospital course for all individuals. Patients who developed AKI after the last specimen collection timepoint were excluded. Controls were defined as individuals who developed AKI stage 1 or did not develop AKI during their hospital course.

### Serum collection and Processing

Blood samples were collected in Serum Separation Tubes (SST) with a polymer gel for serum separation as previously described^35^. Samples were centrifuged at 1200 *g* for 10 minutes at 20°C. After centrifugation, serum was pipetted to a 15 mL conical tube. Serum was then aliquoted into cryovials and stored at −80°C.

### Definition of Acute Kidney Injury

We defined AKI (stage 2 or 3) as per Kidney Disease Improving Global Outcomes (KDIGO) criteria: an increase in serum creatinine of at least 2.0 times the baseline creatinine^36^. For patients with previous serum creatinine measurement available in the 365 days prior to admission, the minimum value in this period was considered the baseline creatinine. For patients without a baseline creatinine in this period, a baseline value was calculated based on an estimated glomerular filtration rate (eGFR) of 75 ml/min per 1.73 m^2^ as per the KDIGO AKI guidelines.

### Clinical data collection

We collected demographic and laboratory data collected as part of standard medical care from an institutional electronic health record (EHR) database. We defined clinical comorbidities using diagnostic codes recorded in the EHR before the current hospital admission. To account for disease severity at the time of specimen collection, we defined supplemental oxygen requirement as 0 if the patient was not receiving supplemental oxygenation or on nasal cannula, 1 if the patient was receiving non-invasive mechanical ventilation (CPAP, BIPAP), or 2 if the person was receiving invasive mechanical ventilation.

### Somalogic proteomic assay

We used the *SomaScan* discovery platform to quantify levels of protein expression. The *SomaScan* platform is a highly multiplexed aptamer based proteomic assay based on Slow Off-rate Modified single-stranded DNA Aptamers (SOMAmers) capable of simultaneously detecting 4497 proteins in biological samples in the form of relative fluorescent units (RFUs). The assay was run using the standard 12 hybridization normalization control sequences to assess for variability in the Agilent plate quantification process, five human calibrator control pooled replicates, and 3 quality control pooled replicates to control for batch effects. Standard preprocessing protocols were applied as per Somalogic’s guidelines published previously^37^ The specificity and stability of the SOMAScan assay has been described previously^38^ Briefly, the data was first normalized using the 12 hybridization controls to remove hybridization variation within a run. Then, median signal normalization is performed with calibrator samples across plates to remove variation in sample-to-sample differences attributable to variations due to pipetting, reagent concentrations, assay timings and other technical aspects. Data was then calibrated to remove assay differences between runs. Standard Somalogic acceptance criteria for quality control metrics were used (plate scale factor between 0.4 and 2.5 and 85% of QC ratios between 0.8 and 1.2). Samples with intrinsic issues such as reddish appearance or low sample volume were also removed as part of the Somalogic quality control protocol. After quality control and normalization procedures, the resulting relative fluorescence unit (RFU) values were log2 transformed.

### Dimensionality reduction

Principal component analysis (PCA) was performed using log2 transformed RFU values of all proteins. Pairwise plots of the top three principal components were plotted.

### Differential expression analysis for prevalent AKI

Using data from the AKI cohort, log_2_ transformed normalized protein values were modelled using multivariable linear regression in the Limma framework^39^ Models were adjusted for age, sex, history of chronic kidney disease (CKD), and supplemental oxygen requirement (0,1, or 2 [see above]) at the time of specimen collection. P-values were adjusted using the Benjamin-Hochberg procedure to control the false discovery rate (FDR) at 5%.

### Proteins associated with change in creatinine

Similarly, we also computed the maximum change in creatinine during hospitalization from baseline for each individual in the discovery cohort. We then associated protein expression values measured at the last available timepoints with maximal change in creatinine to identify a dose response relationship. Models were adjusted for age, sex, history of chronic kidney disease (CKD), and supplemental oxygen requirement (0,1, or 2 [see above]) at the time of specimen collection.

### Proteomic characterization of long-term kidney function in discovery cohort

Outpatient creatinine values measured after discharge were used to compute estimated glomerular filtration rate (eGFR) values the CKD-EPI equation. All values were taken from the EHR as part of routine clinical care with follow-up until 12/2/2021. To determine whether AKI associated protein expression correlated with post-discharge kidney function, we fit a mixed effects linear regression model with random intercept. Using the discovery cohort, protein expression of AKI-associated proteins measured at the last available timepoint during admission was used. The dependent variable was eGFR and the model was adjusted for age, sex, baseline creatinine, history of CKD, maximum AKI stage during the hospital admission, and day of eGFR measurement after hospital discharge. Models included a random effect of patient ID to adjust for correlation between eGFR values taken from the same individuals. Significance was evaluated using a t-test with Satterthwaite degrees of freedom implemented in the lmerTest R package^40^. P values were adjusted using the Benjamin-Hochberg procedure to control the false discovery rate (FDR) at 5%.

We then plotted the post-discharge eGFR values over time for individuals separated by protein expression tertiles (bottom 33rd percentile, middle 33th percentile, and top 33th percentile). We transformed data using the LOESS smoothing function as implemented in the ggplot R package.

### Data analysis and visualization

We performed all statistical analysis using R version 4.0.3. Protein-protein interaction (PPI) network was constructed using the Network X package in Python v3.4.10 to display a Minimum Spanning Tree (MST) using Prim’s algorithm. Network clustering was conducted using the MCL cluster algorithm and functional enrichment was carried out using the STRING^41^ database in Cytoscape^42^. Using results from a recent publication^30^, we also annotated protein quantitative trait loci (pQTLs) for the set of COVID AKI-associated proteins. For each AKI-associated protein, we determined whether or not cis and trans pQTL associations had been reported.

### Data and Code Availability

Data is available by contacting the senior author, Girish Nadkarni.

Code is available at https://github.com/Nadkarni-Lab/aki_covid_proteomics

## RESULTS

### Discovery and Validation Cohort Overview

To discover proteins associated with COVID-AKI, we enrolled a prospective cohort of patients hospitalized with COVID-19 admitted between March 24, 2020 and August 26, 2020 into a biobank as previously described^34^. Cases were defined as patients who developed AKI (stage 2 or 3) during their hospital admission and controls included all other patients (**Fig 1**). Characteristics of cases and controls in the discovery cohort are provided in **Table 1**. Patients who developed AKI were significantly older (67 vs. 60 years, p <0.001), more commonly Hispanic/Latino (48% vs 37%, p =0.012), and had a greater prevalence of atrial fibrillation (17% vs 8%, p =0.008), diabetes (37% vs 20%, p <0.001), and chronic kidney disease (20% vs 4%, p< 0.001). We then validated these associations in an external cohort from Quebec, Canada. Characteristics of the validation cohort are provided in **Supplementary Table 1.** In the validation cohort, compared to controls, AKI (stage 2 or 3) cases had a significantly higher prevalence of CKD (29% vs 11%, p = 0.01) and a higher rate of intubation at the time of blood draw (49% vs 13%).

**Table 1:**
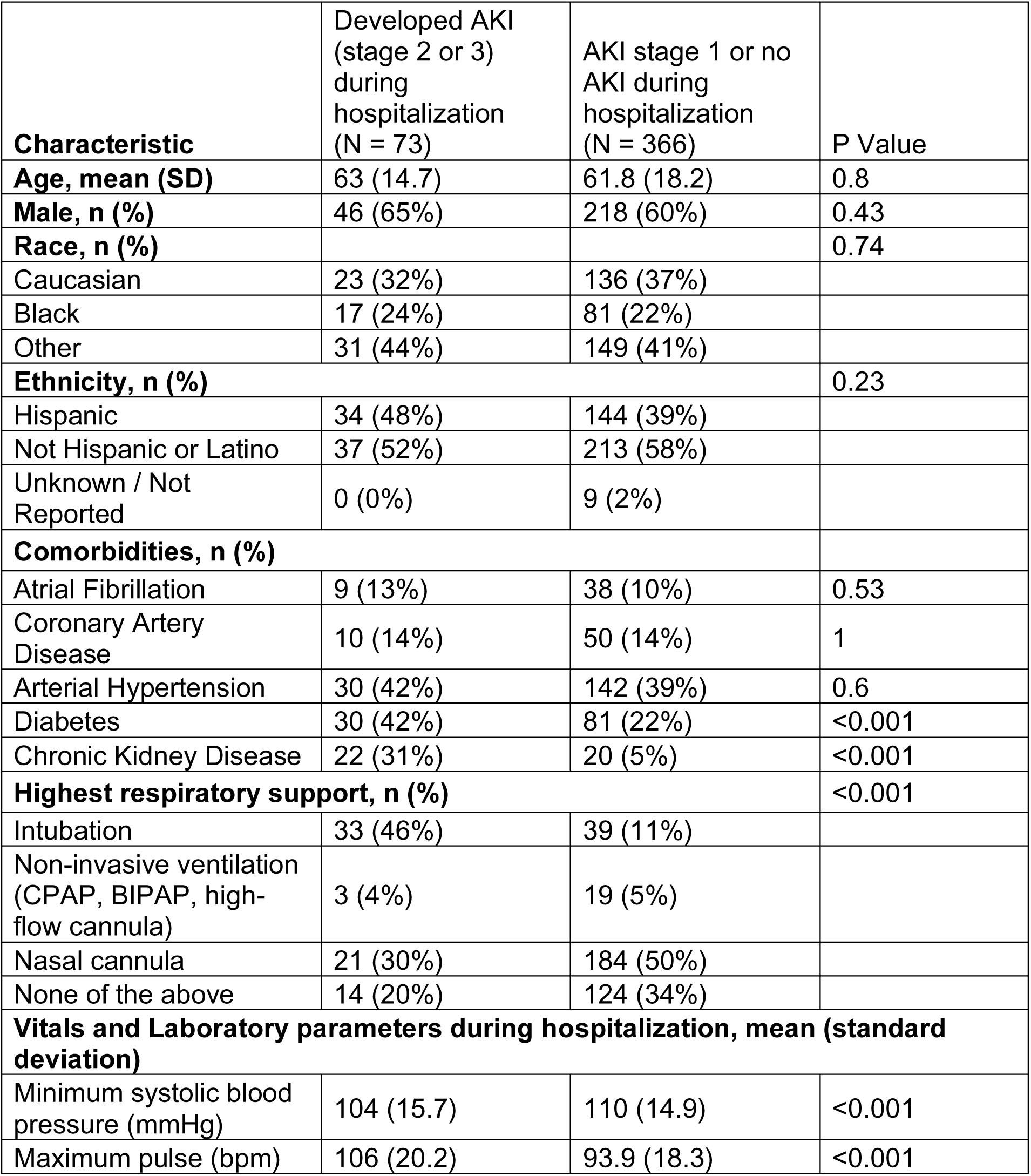
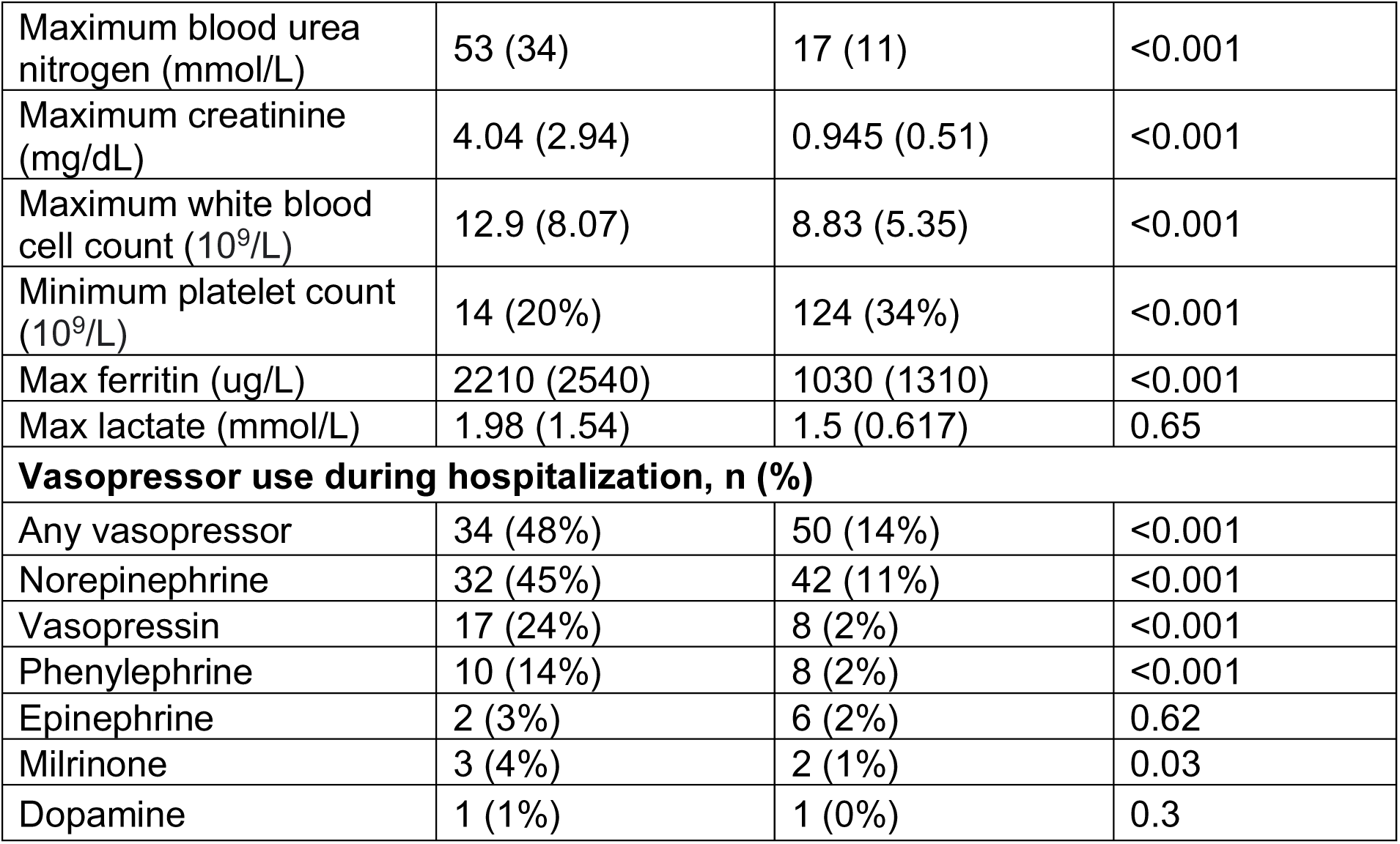
Demographic and clinical comorbidities of patients hospitalized with COVID-19 in the discovery cohort separated by the development of AKI (stage 2 or 3) during their hospital course.

| **Characteristic** | **Developed AKI**<br>**(stage 2 or 3)**<br>**during**<br>**hospitalization**<br>**(N = 73)** | **AKI stage 1 or no**<br>**AKI during**<br>**hospitalization**<br>**(N = 366)** | **P Value** |
| --- | --- | --- | --- |
| **Age, mean (SD)** | 63 (14.7) | 61.8 (18.2) | 0.8 |
| **Male, n (%)** | 46 (65%) | 218 (60%) | 0.43 |
| **Race, n (%)** |  |  | 0.74 |
| Caucasian | 23 (32%) | 136 (37%) |  |
| Black | 17 (24%) | 81 (22%) |  |
| Other | 31 (44%) | 149 (41%) |  |
| **Ethnicity, n (%)** |  |  | 0.23 |
| Hispanic | 34 (48%) | 144 (39%) |  |
| Not Hispanic or Latino | 37 (52%) | 213 (58%) |  |
| Unknown / Not<br>Reported | 0 (0%) | 9 (2%) |  |
| **Comorbidities, n (%)** |  |  |  |
| Atrial Fibrillation | 9 (13%) | 38 (10%) | 0.53 |
| Coronary Artery<br>Disease | 10 (14%) | 50 (14%) | 1 |
| Arterial Hypertension | 30 (42%) | 142 (39%) | 0.6 |
| Diabetes | 30 (42%) | 81 (22%) | <0.001 |
| Chronic Kidney Disease | 22 (31%) | 20 (5%) | <0.001 |
| **Highest respiratory support, n (%)** |  |  | <0.001 |
| Intubation | 33 (46%) | 39 (11%) |  |
| Non-invasive ventilation<br>(CPAP, BIPAP, high-<br>flow cannula) | 3 (4%) | 19 (5%) |  |
| Nasal cannula | 21 (30%) | 184 (50%) |  |
| None of the above | 14 (20%) | 124 (34%) |  |
| **Vitals and Laboratory parameters during hospitalization, mean (standard deviation)** |  |  |  |
| Minimum systolic blood<br>pressure (mmHg) | 104 (15.7) | 110 (14.9) | <0.001 |
| Maximum pulse (bpm) | 106 (20.2) | 93.9 (18.3) | <0.001 |
| Maximum blood urea nitrogen (mmol/L) | 53 (34) | 17 (11) | <0.001 |
| Maximum creatinine (mg/dL) | 4.04 (2.94) | 0.945 (0.51) | <0.001 |
| Maximum white blood cell count (10<sup>9</sup>/L) | 12.9 (8.07) | 8.83 (5.35) | <0.001 |
| Minimum platelet count (10<sup>9</sup>/L) | 14 (20%) | 124 (34%) | <0.001 |
| Max ferritin (ug/L) | 2210 (2540) | 1030 (1310) | <0.001 |
| Max lactate (mmol/L) | 1.98 (1.54) | 1.5 (0.617) | 0.65 |
| **Vasopressor use during hospitalization, n (%)** |  |  |  |
| Any vasopressor | 34 (48%) | 50 (14%) | <0.001 |
| Norepinephrine | 32 (45%) | 42 (11%) | <0.001 |
| Vasopressin | 17 (24%) | 8 (2%) | <0.001 |
| Phenylephrine | 10 (14%) | 8 (2%) | <0.001 |
| Epinephrine | 2 (3%) | 6 (2%) | 0.62 |
| Milrinone | 3 (4%) | 2 (1%) | 0.03 |
| Dopamine | 1 (1%) | 1 (0%) | 0.3 |

### Identification of proteins associated with prevalent AKI

In the discovery cohort, serum levels of 4496 proteins were quantified using the *SomaScan* platform using samples collected at multiple timepoints during the hospital course (**Supplementary Table 2**) as previously described^34^. We first identified proteins associated with prevalent AKI using measurements taken after the onset of AKI in cases and the last available measurement in controls (**Fig 1**, 71 cases and 366 controls). The top three principal components (PCs) distinctly separate samples by case status (**Fig 2A**). We fit a multivariable linear regression model for the log2 normalized protein expressions adjusted for age, sex, history of chronic kidney disease (CKD), and maximum oxygen requirement at the time of blood draw. We identified 413 proteins with higher plasma abundances and 30 proteins with lower plasma abundance (**Supplementary Table 3**).

**Fig 2:**
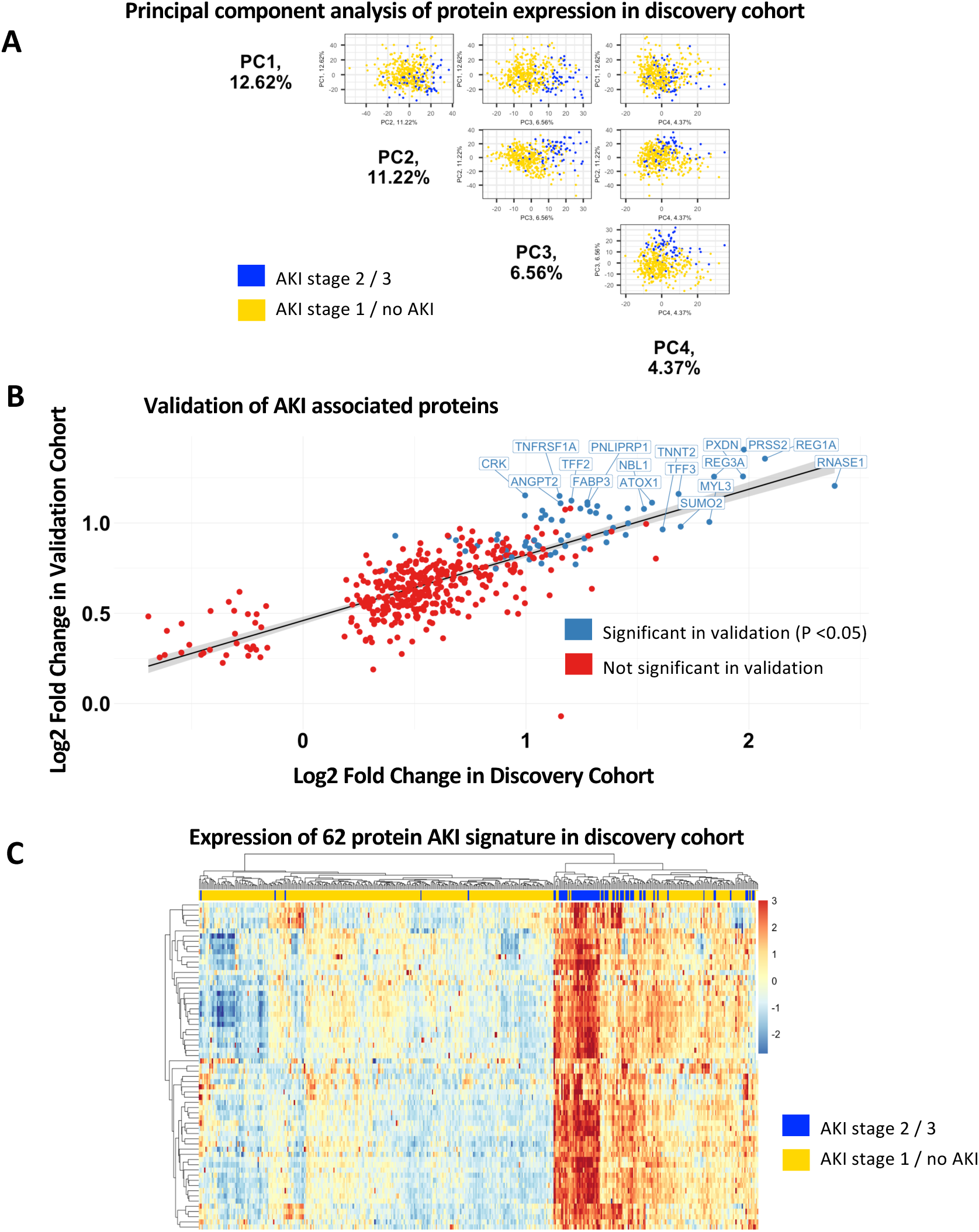
**A.** Top 3 Principal Components show separation of the sample by AKI (stage 2 or 3) case status. **B.** External validation of AKI associated proteins in the discovery cohort shows high correlation with increased risk of AKI with significance of p<0.05. **C.** Expression heatmap shows a distinct separation of the cases and controls using the 62 significant proteins identified from the validation cohort in the discovery cohort.

### Validation of AKI-associated proteins

We then performed an external validation of AKI-associated proteins in a prospective biobank cohort from Quebec, Canada. Of the 443 proteins significantly associated with AKI in the discovery cohort (FDR adjusted P<0.05), 62 were also associated with AKI in the validation cohort (p <0.05, **Table 2**). All validated proteins were associated with an increased risk of AKI. The fold changes of validated proteins in the discovery and validation cohort were highly correlated with a Pearson correlation of 0.71 (**Fig 2B**). The 62-protein signature distinctly separated AKI cases from cohorts in the discovery cohort (**Fig 2C**). Using reported plasma pQTL associations from a recent publication, of the 62 AKI associated proteins, 45 had both cis and trans pQTLS, 14 had only trans pQTLs, and 2 had cis pQTLs (Supplementary Figure 1). Protein-protein interaction (PPI) network analysis revealed enrichment of several highly connected proteins, including LCN2 (alternative name: NGAL), REG3A, and MB (**Fig 4A**). The AKI-associated protein network also included a cluster of cardiac structural proteins (**Fig 4B**), TNNT2, TTN, MYL3, SRL, and NPPB (alternative name: BNP).

**Table 2:** Validation of AKI-associated proteins. Strength of association of proteins significantly associated with AKI (stage 2 or 3) in both the discovery (adjusted P <0.05) and validation cohort (P <0.05) are provided. Significance was determined by fitting a linear model adjusted for age, sex, history of CKD, and maximum oxygen requirement at the time of blood draw.

| Gene Name | Gene Symbol | Discovery cohort |  |  |  | Validation cohort |  |  |
| --- | --- | --- | --- | --- | --- | --- | --- | --- |
|  |  | logF C | Fold Change | P | FDR | LogF C | Fold Change | P |
| Trypsin-2 | PRSS2 | 1.98 | 3.94 | 3.16E-26 | 1.42E-22 | 1.41 | 2.65 | 0.0019 |
| Lithostathine-1-alpha | REG1A | 2.07 | 4.20 | 9.23E-38 | 4.15E-34 | 1.36 | 2.56 | 0.0076 |
| Peroxidasin homolog | PXDN | 1.97 | 3.93 | 1.39E-34 | 6.25E-31 | 1.26 | 2.39 | 0.0047 |
| Regenerating islet-derived protein 3-alpha | REG3A | 1.84 | 3.59 | 6.44E-26 | 2.90E-22 | 1.26 | 2.39 | 0.0139 |
| Ribonuclease pancreatic | RNASE1 | 2.38 | 5.22 | 1.53E-38 | 6.87E-35 | 1.21 | 2.31 | 0.0021 |
| Trefoil factor 3 | TFF3 | 1.68 | 3.21 | 2.13E-41 | 9.55E-38 | 1.16 | 2.24 | 0.0238 |
| Adapter molecule crk | CRK | 1.00 | 1.99 | 8.67E-18 | 3.90E-14 | 1.15 | 2.22 | 0.0219 |
| Tumor necrosis factor receptor superfamily member 1A | TNFRSF1A | 1.15 | 2.22 | 3.86E-21 | 1.74E-17 | 1.15 | 2.22 | 0.0281 |
| Trefoil factor 2 | TFF2 | 1.20 | 2.30 | 3.36E-16 | 1.51E-12 | 1.12 | 2.18 | 0.0273 |
| Fatty acid-binding protein, heart | FABP3 | 1.28 | 2.42 | 3.09E-21 | 1.39E-17 | 1.11 | 2.17 | 0.0356 |
| Copper transport protein ATOX1 | ATOX1 | 1.57 | 2.96 | 2.04E-40 | 9.16E-37 | 1.11 | 2.16 | 0.0072 |
| Angiopietin-2 | ANGPT2 | 1.15 | 2.23 | 1.44E-08 | 6.45E-05 | 1.11 | 2.16 | 0.0342 |
| Inactive pancreatic lipase-related protein 1 | PNLIPRP1 | 1.28 | 2.42 | 5.64E-15 | 2.54E-11 | 1.10 | 2.15 | 0.0296 |
| Trypsin-3 | PRSS3 | 1.32 | 2.50 | 9.07E-18 | 4.08E-14 | 1.09 | 2.13 | 0.0069 |
| Acyl-CoA-binding protein | DBI | 1.46 | 2.74 | 1.82E-44 | 8.20E-41 | 1.08 | 2.12 | 0.0360 |
| Ganglioside GM2 activator | GM2A | 1.24 | 2.37 | 2.02E-40 | 9.09E-37 | 1.08 | 2.11 | 0.0088 |
| Neuroblastoma suppressor of tumorigenicity 1 | NBL1 | 1.53 | 2.89 | 1.79E-37 | 8.03E-34 | 1.08 | 2.11 | 0.0099 |
| Apolipoprotein F | APOF | 1.07 | 2.10 | 2.97E-14 | 1.34E-10 | 1.07 | 2.10 | 0.0317 |
| Collagen alpha-1(XXVIII) chain | COL28A1 | 1.29 | 2.44 | 2.29E-34 | 1.03E-30 | 1.06 | 2.09 | 0.0252 |
| Carbonic anhydrase 3 | CA3 | 1.31 | 2.49 | 2.13E-13 | 9.58E-10 | 1.06 | 2.08 | 0.0174 |
| CD59 glycoprotein | CD59 | 1.09 | 2.12 | 5.67E-21 | 2.55E-17 | 1.04 | 2.06 | 0.0318 |
| Ephrin-B2 | EFNB2 | 0.99 | 1.99 | 1.45E-14 | 6.52E-11 | 1.04 | 2.06 | 0.0437 |
| Transmembrane emp24 domain-containing protein 10 | TMED10 | 1.37 | 2.59 | 2.39E-42 | 1.07E-38 | 1.03 | 2.05 | 0.0078 |
| Trypsin-1 | PRSS1 | 1.07 | 2.10 | 3.15E-13 | 1.42E-09 | 1.03 | 2.04 | 0.0456 |
| Cystatin-D | CST5 | 1.11 | 2.16 | 4.02E-13 | 1.81E-09 | 1.02 | 2.03 | 0.0464 |
| Transgelin | TAGLN | 1.16 | 2.24 | 1.60E-27 | 7.20E-24 | 1.01 | 2.02 | 0.0379 |
| Myosin light chain 3 | MYL3 | 1.82 | 3.54 | 2.12E-15 | 9.54E-12 | 1.01 | 2.01 | 0.0127 |
| IGF-like family receptor 1 | IGFLR1 | 1.46 | 2.75 | 1.97E-26 | 8.85E-23 | 1.00 | 2.00 | 0.0242 |
| Small ubiquitin-related modifier 2 | SUMO2 | 1.69 | 3.24 | 2.07E-44 | 9.29E-41 | 0.98 | 1.97 | 0.0469 |
| Tyrosine-protein kinase transmembrane receptor ROR2 | ROR2 | 1.39 | 2.62 | 1.29E-32 | 5.82E-29 | 0.97 | 1.96 | 0.0410 |
| Troponin T, cardiac muscle | TNNT2 | 1.61 | 3.06 | 6.66E-27 | 2.99E-23 | 0.96 | 1.95 | 0.0233 |
| Complexin-2 | CPLX2 | 1.36 | 2.57 | 5.43E-30 | 2.44E-26 | 0.94 | 1.92 | 0.0145 |
| Serine protease inhibitor Kazal-type 7 | SPINK7 | 0.87 | 1.83 | 4.06E-18 | 1.82E-14 | 0.94 | 1.91 | 0.0242 |
| Myoglobin | MB | 1.45 | 2.73 | 7.19E-23 | 3.23E-19 | 0.93 | 1.91 | 0.0427 |
| Cystatin-C | CST3 | 1.19 | 2.28 | 2.09E-41 | 9.38E-38 | 0.93 | 1.91 | 0.0297 |
| Hepatoma-derived growth factor | HDGF | 0.41 | 1.33 | 4.05E-07 | 1.82E-03 | 0.93 | 1.90 | 0.0241 |
| Ig gamma-4, Kappa | IGHG4 | 0.65 | 1.57 | 2.26E-12 | 1.02E-08 | 0.92 | 1.90 | 0.0217 |
| Desmocollin-2 | DSC2 | 1.26 | 2.40 | 2.84E-39 | 1.28E-35 | 0.91 | 1.88 | 0.0249 |
| PILR alpha-associated neural protein | PIANP | 0.87 | 1.83 | 1.47E-24 | 6.60E-21 | 0.91 | 1.88 | 0.0226 |
| Ephrin type-A receptor 2 | EPHA2 | 1.04 | 2.05 | 1.72E-21 | 7.74E-18 | 0.91 | 1.87 | 0.0294 |
| Natriuretic peptides B | NPPB | 0.69 | 1.61 | 8.76E-09 | 3.94E-05 | 0.90 | 1.87 | 0.0399 |
| Chymotrypsinogen B | CTRB1 | 1.00 | 2.00 | 1.62E-09 | 7.29E-06 | 0.90 | 1.86 | 0.0202 |
| Secretogranin-1 | CHGB | 1.36 | 2.57 | 2.28E-37 | 1.02E-33 | 0.89 | 1.86 | 0.0180 |
| Ephrin-B1 | EFNB1 | 0.89 | 1.86 | 1.30E-38 | 5.86E-35 | 0.89 | 1.85 | 0.0365 |
| Parathyroid hormone | PTH | 1.05 | 2.07 | 2.68E-14 | 1.20E-10 | 0.88 | 1.84 | 0.0443 |
| BolA-like protein 1 | BOLA1 | 0.78 | 1.72 | 1.61E-29 | 7.26E-26 | 0.87 | 1.83 | 0.0288 |
| Serine/threonine-protein kinase receptor R3 | ACVRL1 | 1.00 | 1.99 | 9.51E-32 | 4.28E-28 | 0.87 | 1.83 | 0.0200 |
| Tumor necrosis factor receptor superfamily member 19 | TNFRSF19 | 1.06 | 2.08 | 1.84E-33 | 8.29E-30 | 0.87 | 1.83 | 0.0272 |
| Ribonuclease K6 | RNASE6 | 1.18 | 2.26 | 9.28E-29 | 4.17E-25 | 0.87 | 1.83 | 0.0498 |
| Selenoprotein W | SEPW1 | 1.26 | 2.40 | 1.56E-15 | 7.01E-12 | 0.86 | 1.82 | 0.0489 |
| Protein delta homolog 2 | DLK2 | 1.02 | 2.02 | 2.29E-15 | 1.03E-11 | 0.86 | 1.81 | 0.0415 |
| Asialoglycoprotein receptor 1 | ASGR1 | 0.91 | 1.88 | 5.36E-13 | 2.41E-09 | 0.85 | 1.80 | 0.0434 |
| Matrix-remodeling-associated protein 7 | MXRA7 | 0.73 | 1.66 | 7.02E-22 | 3.15E-18 | 0.84 | 1.80 | 0.0288 |
| Neutrophil gelatinase-associated lipocalin | LCN2 | 1.13 | 2.19 | 6.27E-22 | 2.82E-18 | 0.84 | 1.79 | 0.0363 |
| Osteopontin | SPP1 | 1.11 | 2.16 | 4.47E-21 | 2.01E-17 | 0.82 | 1.77 | 0.0370 |
| Protocadherin gamma-A10 | PCDHGA10 | 0.90 | 1.87 | 5.70E-16 | 2.56E-12 | 0.81 | 1.76 | 0.0406 |
| DnaJ homolog subfamily B member 12 | DNAJB12 | 0.94 | 1.91 | 9.54E-17 | 4.29E-13 | 0.81 | 1.75 | 0.0466 |
| Sarcalumenin | SRL | 1.02 | 2.02 | 6.00E-38 | 2.70E-34 | 0.80 | 1.74 | 0.0475 |
| Discoidin domain-containing receptor 2 | DDR2 | 1.07 | 2.10 | 3.40E-21 | 1.53E-17 | 0.78 | 1.71 | 0.0359 |
| Titin | TTN | 1.22 | 2.33 | 5.42E-25 | 2.44E-21 | 0.77 | 1.71 | 0.0281 |
| Colipase | CLPS | 0.86 | 1.82 | 1.95E-17 | 8.77E-14 | 0.75 | 1.68 | 0.0362 |
| Stathmin-3 | STMN3 | 0.37 | 1.29 | 6.95E-09 | 3.13E-05 | 0.74 | 1.67 | 0.0480 |

### Proteins associated with change in creatinine

We then tried to identify which AKI-associated proteins demonstrated a dose dependent relationship with change in creatinine. Using the discovery dataset, we fit a multivariable linear regression model for maximal change in creatinine during hospitalization and found that all 62 AKI-associated proteins were also significantly associated with maximal change in creatinine (P <0.05, Supplementary Table 4).

### Proteomic characterization of post-acute kidney dysfunction

Given the previously reported association of COVID-19 AKI with long-term eGFR decline^43^, we hypothesized that significant proteomic markers associated with COVID-19 AKI are also associated with post-discharge eGFR. We included all outpatient eGFR measurements taken after discharge from patients in the Mount Sinai biobank cohort Of the 437 patients in the cohort, 181 patients had at least one outpatient post-discharge eGFR measurement. The median number of eGFR measurements was 4 with an interquartile of 9. The first post-discharge eGFR was measured at a median of 37 days after discharge. The last post-discharge eGFR was measured at a median of 374 days after discharge (Supplementary Figure 2).

We used a mixed effects linear model accounting for baseline creatinine, AKI stage during the COVID admission and repeated eGFR measurements to associate the 62 protein AKI signature with long-term eGFR. Of the 62 AKI-associated proteins, 25 were significantly (FDR adjusted P <0.05) associated with long-term post-discharge eGFR (**Fig 3, Fig 5A**). All 25 eGFR-associated proteins were negatively correlated with post-discharge eGFR (**Table 3**). However, the strength of association with AKI was not significantly associated with the strength of association with post-discharge eGFR. Proteins most strongly associated (by P value) with decreased post-discharge eGFR included desmocollin-2, trefoil factor 3, transmembrane emp24 domain-containing protein 10, and cystatin-C (**Fig 5B**).

**Table 3:** Association of AKI-associated proteins with post-discharge eGFR. A mixed effects linear model adjusted for age, sex, history of CKD, baseline creatinine, AKI stage during the COVID admission and number repeated eGFR measurements was fit. Proteins significantly associated with eGFR (adjusted P <0.05) are provided.

| **Gene name** | **Coefficient** | **P Value** | **Adjusted P Value** | **Uniprot ID** |
| --- | --- | --- | --- | --- |
| BolA-like protein 1 | -18.022031 | 1.38x10<sup>-5</sup> | 0.0008 | Q9Y3E2 |
| Cystatin-C | -17.63355 | 3.40x10<sup>-7</sup> | 2.14x10<sup>-5</sup> | P01034 |
| Desmocollin-2 | -16.597692 | 1.63x10<sup>-9</sup> | 1.03 x 10<sup>-7</sup> | Q02487 |
| Transmembrane emp24 domain-containing protein 10 | -15.819807 | 1.36x10<sup>-8</sup> | 8.54x10<sup>-7</sup> | P49755 |
| Ephrin-B1 | -15.167647 | 0.0002 | 0.01 | P98172 |
| Serine/threonine-protein kinase receptor R3 | -13.939695 | 7.64x10<sup>-5</sup> | 0.004 | P37023 |
| Ganglioside GM2 activator | -13.060806 | 1.17x10<sup>-5</sup> | 0.0007 | P17900 |
| Matrix-remodeling-associated protein 7 | -11.88046 | 0.0002 | 0.017 | P84157 |
| PILR alpha-associated neural protein | -11.803896 | 3.23x10<sup>-5</sup> | 0.002 | Q8IYJ0 |
| Trefoil factor 3 | -11.738116 | 1.60x10<sup>-8</sup> | 1.01x10<sup>-6</sup> | Q07654 |
| Copper transport protein ATOX1 | -11.216693 | 4.38x10<sup>-6</sup> | 0.0002 | O00244 |
| Serine protease inhibitor Kazal-type 7 | -11.157036 | 5.77x10<sup>-6</sup> | 0.0003 | P58062 |
| IGF-like family receptor 1 | -11.007485 | 4.79x10<sup>-7</sup> | 3.02x10<sup>-5</sup> | Q9H665 |
| Complexin-2 | -10.73936 | 1.33x10<sup>-6</sup> | 8.36 x 10<sup>-5</sup> | Q6PUV4 |
| Collagen alpha-1(XXVIII) chain | -10.047217 | 0.0001 | 0.006 | Q2UY09 |
| CD59 glycoprotein | -9.9917438 | 1.74x10<sup>-5</sup> | 0.001 | P13987 |
| Ribonuclease K6 | -9.9898356 | 0.0001 | 0.006 | Q93091 |
| Transgelin | -9.9393974 | 0.0001 | 0.01 | Q01995 |
| DnaJ homolog subfamily B member 12 | -8.2638876 | 0.0004 | 0.03 | Q9NXW2 |
| Protein delta homolog 2 | -7.7406286 | 0.0002 | 0.01 | Q6UY11 |
| Discoidin domain-containing receptor 2 | -7.6524963 | 0.0006 | 0.04 | Q16832 |
| Ribonuclease pancreatic | -7.40997 | 8.01x10<sup>-7</sup> | 5.05x10<sup>-5</sup> | P07998 |
| Ephrin-B2 | -7.2943084 | 0.0001 | 0.009 | P52799 |
| Peroxidasin homolog | -7.2282085 | 1.75x10<sup>-5</sup> | 0.001 | Q92626 |
| Trefoil factor 2 | -7.2179491 | 1.2x10<sup>-5</sup> | 0.0007 | Q03403 |

**Fig 3:**
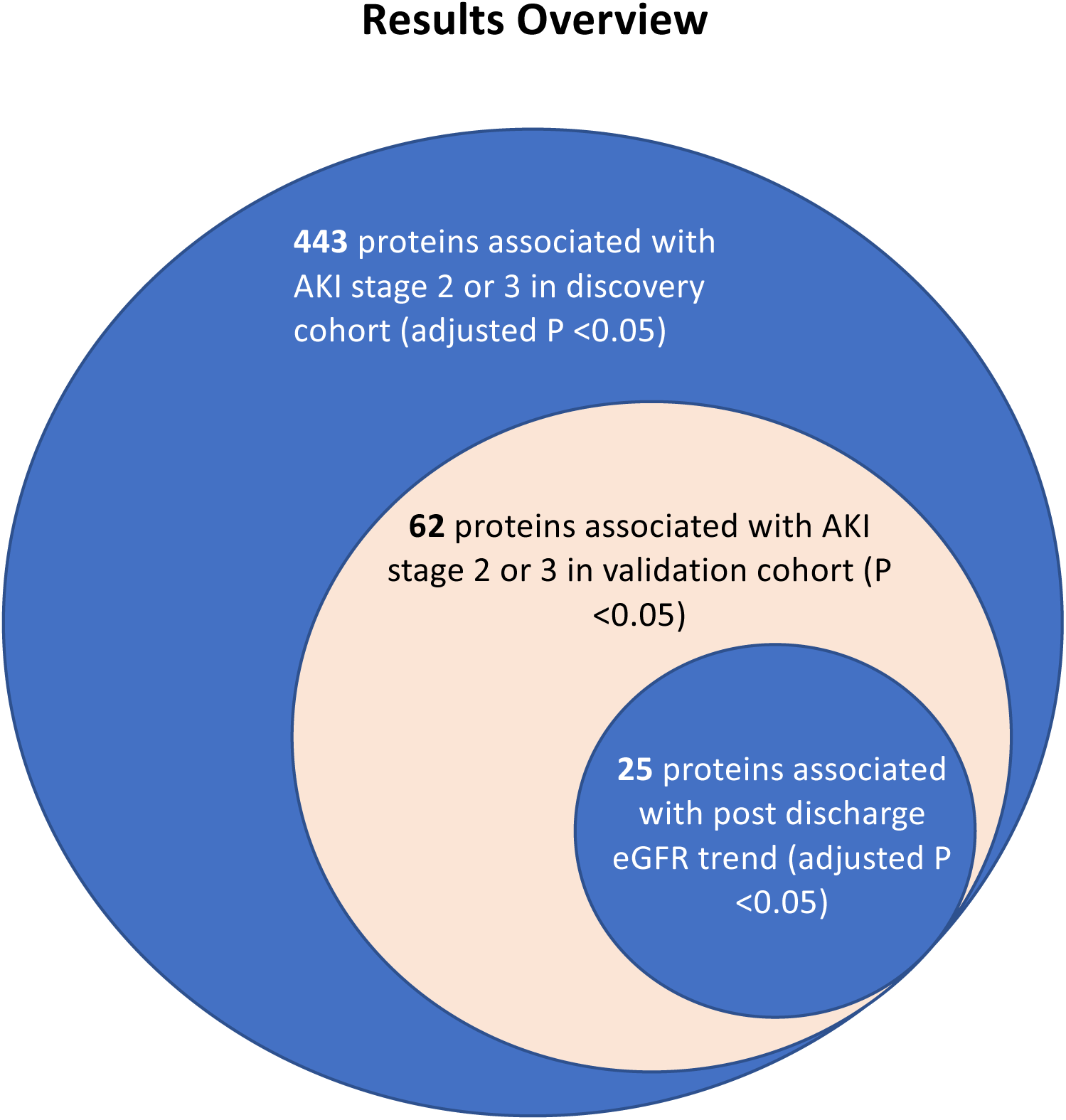
Nested Venn diagram of the analyses performed.

**Fig 4:**
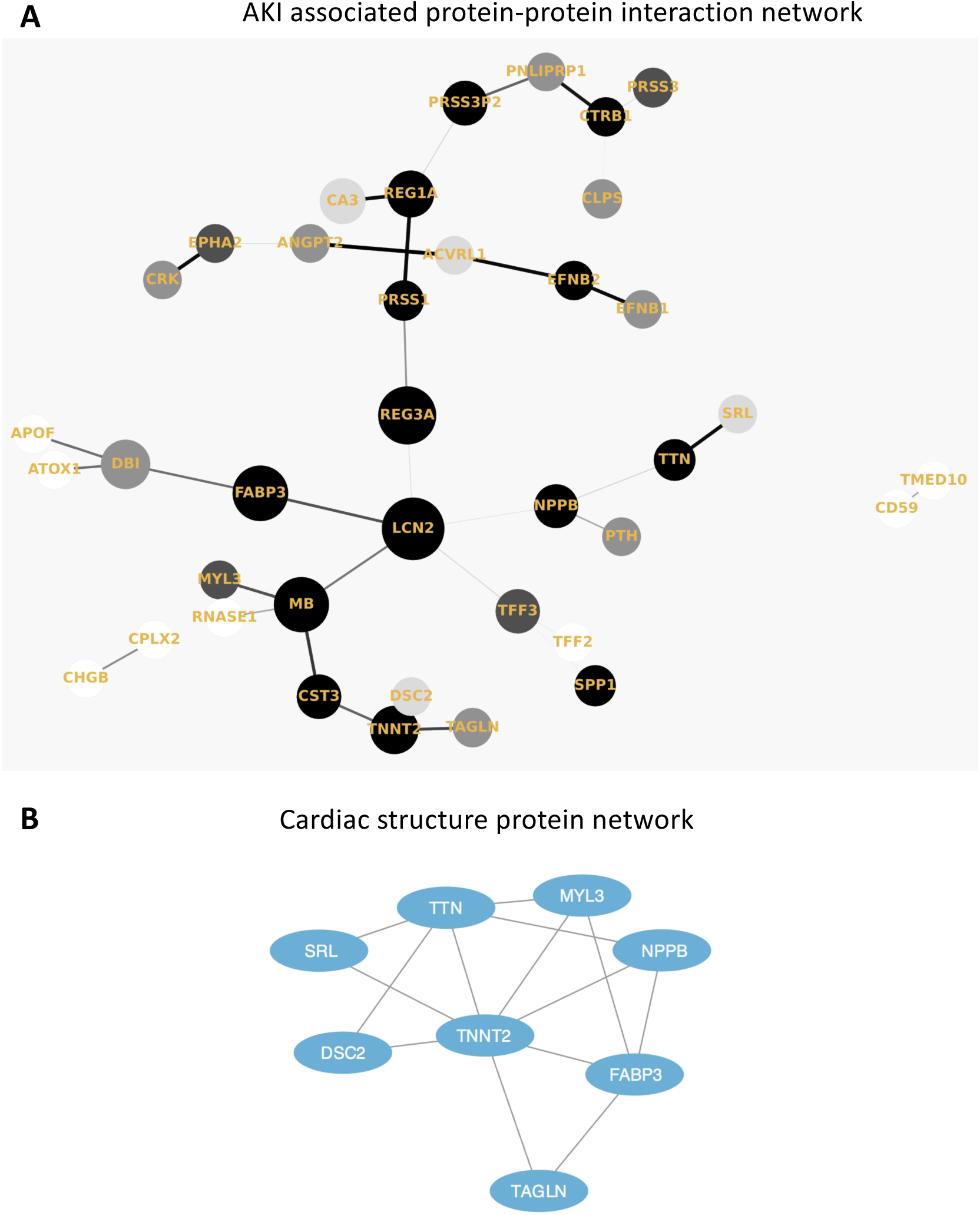
A. Protein–protein interaction (PPI) network (Minimum Spanning Tree) of the 62 overlapping AKI associated proteins with a score >0.4. The size of each node corresponds to number of interactions and the thickness of the edges represent the weight of the interactions between the nodes. B. MCL algorithm was used to identify tightly connected cluster of proteins which was functionally enriched for cardiac structure proteins using the STRING database.

**Fig 5:**
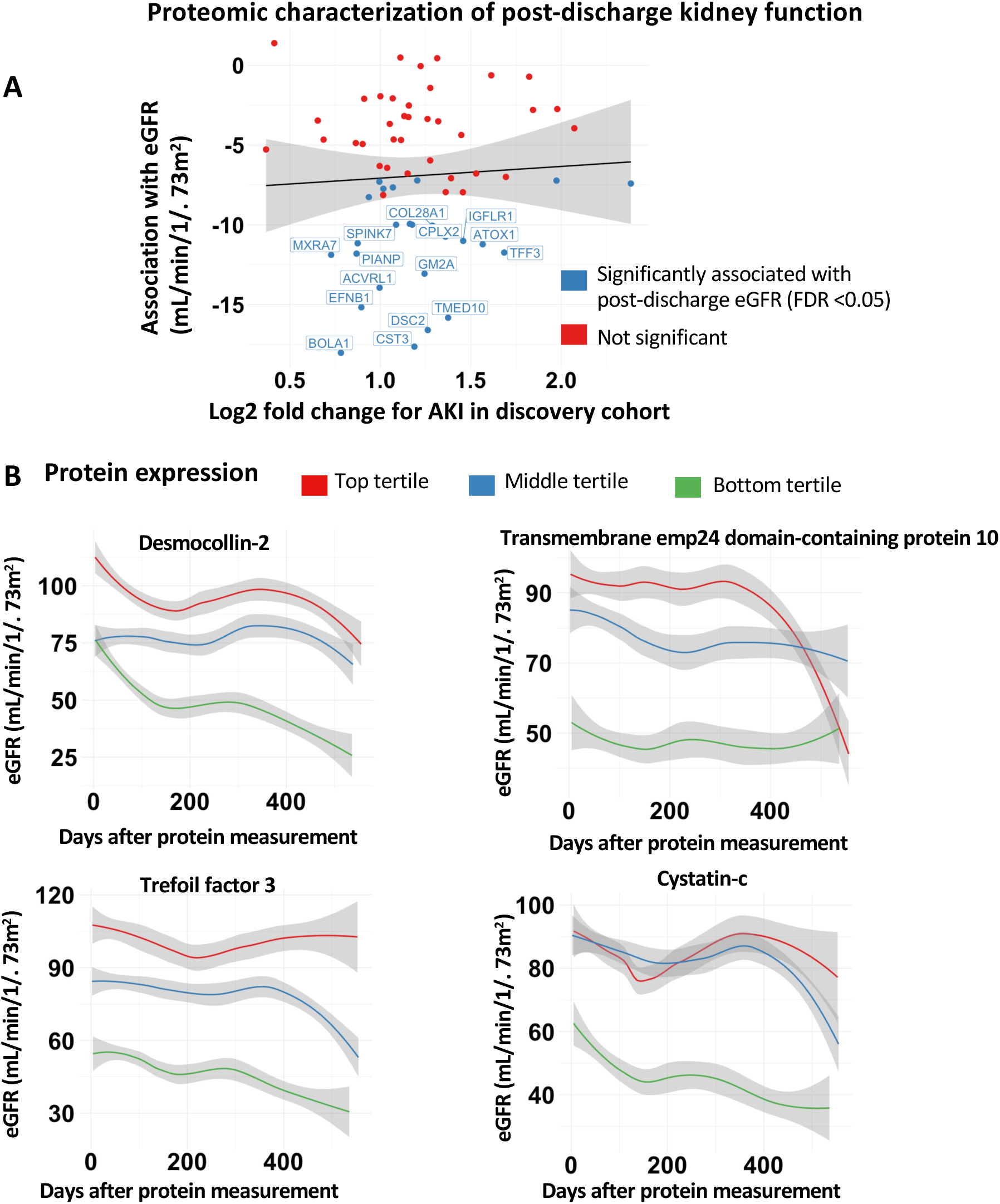
Proteomic characterization of long-term eGFR decline. A. Comparison of strengths of association with AKI and long term eGFR for proteins associated with AKI in both the discovery and validation cohorts. B. Trend in eGFR values separated by protein expression for tertiles for proteins most significantly (by P value) associated with eGFR trend.

## DISCUSSION

Using proteomic profiling in two large groups of patients hospitalized with COVID-19, we report several observations. First, we identified specific protein markers of AKI and post-discharge kidney dysfunction, both well-documented sequelae of COVID-19^4, 43^. Second, in the acute phase, tubular injury and hemodynamic perturbation may play a role. Thus, characterization of the peripheral blood suggests specific large-scale perturbations of the proteome that accompany both AKI and long-term eGFR decline with implications for more specific prognostic models and targeted therapeutic development.

Based on our results, we hypothesize that COVID AKI may involve several mechanisms: tubular injury, neutrophil activation, and hemodynamic perturbation. First, we found significantly higher plasma abundances of NGAL (LCN2), a canonical marker of tubular injury that is also involved in neutrophil activation. NGAL is secreted by circulating neutrophils and kidney tubular epithelium in response to systemic inflammation or ischemia. Since renal tubular epithelial cells express the angiotensin-converting enzyme 2 (ACE2) receptor which enables SARS-CoV2 viral entry into cells, direct tubular infection may cause the release of NGAL into the serum and urine. This potential mechanism is supported by our results and remains a testable hypothesis. Although NGAL is a known marker for intrinsic AKI accompanied by tubular injury, it is relatively insensitive to pre-renal AKI caused by hemodynamic disturbance^44, 45^. However, our results demonstrate higher plasma abundance of BNP, a protein released in the setting of volume overload as well as several cardiac structural proteins (cardiac troponin T, titin, myosin light chain 1, and sarcalumenin). This proteomic signature may represent an impaired crosstalk between the cardiovascular system and kidney in which myocardial injury leads to decreased renal perfusion and eventual AKI. Myocardial injury has been previously reported in patients hospitalized with COVID-19^6^ and thus may contribute to the multifactorial nature of COVID-AKI. It is worth nothing that in addition to myocardial injury, BNP may also be increased in critical illness due to pro-inflammatory cytokine release.

Since COVID-AKI increases the risk of long-term eGFR decline^43^, we then sought to determine whether these two phenomena shared common proteomic markers. Surprisingly, we found that although almost half of the AKI-associated proteins were also significantly associated with post-discharge eGFR decline, the strengths of associations were not correlated. While COVID-AKI is likely caused by a combination of intrinsic tubular injury and hemodynamic disturbance in the setting of critical illness, long term eGFR decline was associated with increased expression of trefoil factor 3 (TFF3), a known prognostic marker for incident CKD^46^. Trefoil factors are a class of small peptides expressed in colonic and urinary tract epithelia that play essential roles in regeneration and repair of epithelial tissue^47, 48^. Immunohistochemistry reveals TFF3 expression is localized to the tubular epithelial cells in kidney specimens from patients with CKD^46^, suggesting that long term eGFR decline may be associated with renal tubular epithelial damage. The exact pathological role of TFF3 in the renal tubules is unclear but it has been hypothesized to play a role in repair of kidney damage^49^. Additionally, TFF3 release from the renal interstitium has also been hypothesized to direct the epithelial-to-mesenchymal transition (EMT) in renal interstitial fibrosis, a main pathway that leads to ESKD^46^. Our results implicate tubular damage in both AKI and long term eGFR decline suggesting that SARS-CoV2 may preferentially target this region of the nephron. While AKI in the acute setting may be a result of ischemia and decreased renal perfusion associated with critical illness, the specific elevation of TFF3 associated with eGFR decline implicates a more general pattern of tubular injury that underlies COVID mediated kidney dysfunction. Since the ACE2, is preferentially expressed in the tubular epithelial cells of the kidney^50, 51^, the elevation of markers of tubular damage in the plasma may represent direct viral invasion of tubular epithelia cells. However, again this would need to be tested using biopsy/autopsy specimens or other mechanistic studies. Direct viral entry into the kidney remains controversial and using our current data we are not able to comment on this mechanism.

Our study should be interpreted in the context of certain limitations. First, samples were collected during the hospital course of patients with confirmed COVID-19. However, the timepoints were not systematic due to logistical challenges during the peak of the COVID-19 pandemic and thus are not standardized between patients. Since a subset of patients had AKI at the time of admission, these patients were excluded from our analysis since specimens were collected after admission. Additionally, we did not include patients who developed AKI without COVID and were unable to determine whether COVID-AKI has unique proteomic markers compared to other forms of sepsis-AKI. Thus, our AKI cases may be biased towards less severe presentations. Second, since kidney injury is usually not an isolated phenomenon in critically ill patients, the protein expression changes observed may have been partially due to damage to other organs, such as the lung, liver, and heart. However, we accounted for non-kidney damage by adjusting for the highest level of ventilatory support and thus our results are likely a reflection of kidney injury. However, our results do show the importance of crosstalk between the cardiac system and the kidneys. Additionally, we did not include proteomic measurements from urine specimens and thus it is unclear whether poor filtration or resorption of proteins plays a role in peripheral blood protein concentrations. For example, poor resorption of cystatin-C in the setting of AKI may have led to the increased peripheral blood cystatin-C that we report. Our study was adequately powered to detect effect sizes of greater than or equal to 1.6. Additionally, since we enrolled patients only from March-October 2020, we cannot generalize our findings to other COVID-19 variants and time periods. Although we adjusted our regression models for history of CKD, it is possible that unmeasured confounding due to preexisting impaired kidney function has not completely been controlled our in our analyses. Finally, our cohort did not include autopsy or kidney biopsy specimens. Histopathological analysis of kidney specimens is necessary to determine the mechanism of AKI and whether viral particles are present in the kidney.

In conclusion, we provide the first comprehensive characterization of the plasma proteome of AKI and long term eGFR decline in hospitalized COVID-19 patients. Our results suggest in the setting of COVID-AKI and post-discharge kidney dysfunction there is evidence of tubular damage in the peripheral blood but that in the acute setting, several factors including hemodynamic disturbance and myocardial injury also play a role.

## Data Availability

Data is available by contacting the senior author, Girish Nadkarni.
Code is available at https://github.com/Nadkarni-Lab/aki_covid_proteomics

## Author contributions

Conceptualization: IP, GNN, AWC, SZ, NB, ES, EK, LC, SC, BM, SD

Methodology: IP, PJ, RT, KC, SA, AV, SJ, MP, RO, SK, SZ, EUA, FFG, MSF

Investigation: IP, PJ, RT, EK, SZ, SJ, KC, AK, SG, HX, JH, MP, KA, CB, KN, RO

Visualization: IP, PJ

Funding acquisition: GNN, AWC, BSG, MM, NB

Supervision: GNN, AWC, BSG, MM, SK, NB, ES, JCH

Writing: IP, PJ, GNN, SK

## Acknowledgements

We would like to thank the scientists at Somalogic for their assistance with technical and scientific questions about the assay.

