## Supplementary for "Proteomic Characterization of Acute Kidney Injury in Patients Hospitalized with SARS-CoV2 Infection"

### Supplementary Material

**Supplementary Table 1**: Demographic and clinical comorbidities of patients in the validation cohort. Cases include patients who developed AKI stage 2 or 3 during hospitalization. Controls include patients who developed AKI stage 1 or did not develop AKI during hospitalizaiton.

| **Characteristic** | **Developed AKI (stage 2 or 3) during hospitalization (N = 35)** | **AKI stage 1 or no AKI during hospitalization (N = 226)** | **P Value** |
|---|---|---|---|
| **Age, mean (SD)** | 68.4 (17.1) | 70.2 (17.5) | 0.534 |
| **Male, n (%)** | 14 (40%) | 120 (53%) | 0.203 |
| **Ancestry, n (%)** | | | 0.341 |
| European | 22 (63%) | 151 (67%) | |
| East Asian | 2 (6%) | 22 (10%) | |
| African | 5 (14%) | 27 (12%) | |
| South Asian | 1 (3%) | 4(2%) | |
| Mixed American | 1 (3%) | 4(2%) | |
| Unknown | 1 (3%) | 0 (0%) | |
| Other | 3 (9%) | 18 (8%) | |
| **Comorbidities, n (%)** | | | |
| Atrial Fibrillation | 5 (14%) | 38 (17%) | 0.811 |
| Coronary Artery Disease | 9 (26%) | 32 (14%) | 0.0859 |
| Arterial Hypertension | 23 (66%) | 142 (63%) | 0.851 |
| Diabetes | 15 (43%) | 77 (34%) | 0.344 |
| Chronic Kidney Disease | 10 (29%) | 25 (11%) | 0.0130 |
| **Highest respiratory support, n (%)** | | | <0.001 |
| Intubation | 17 (49%) | 30 (13%) | |
| Non-invasive ventilation (CPAP, BIPAP, high-flow cannula) | 3 (9%) | 23 (10%) | |
| Nasal cannula | 5 (14%) | 94 (42%) | |
| None of the above | 10 (29%) | 79 (35%) | |

**Supplementary Table 2**: Distribution of the number of SomaScan timepoints per person in the discovery cohort.

| **Number of Somascan Time points** | **Number of patients** |
|---:|---:|
| 1 | 203 |
| 2 | 137 |
| 3 | 69 |
| 4 | 30 |
| 5 | 15 |
| 6 | 1 |
| 10 | 1 |

**Supplementary Table 3:**

Proteins that were significantly associated with AKI stage 2 or 3 (adjusted P <0.05) in the discovery cohort are provided. Statistical significance was estimated by fitting a linear model adjusted for age, sex, history of CKD, and maximum oxygen requirement at the time of blood draw using the Limma R package[53]. Protein expression data was log2 transformed. P values were adjusted for multiple comparisons using the Bonferroni correction.

For Supplementary Table 3 please see attachment.

**Supplementary Table 4:**

Association of AKI-associated proteins with maximal change in creatinine during hospitalization in the discovery cohort. Maximal change in creatinine was computed with respect to the baseline creatinine for each patient. Statistical significance was estimated by fitting a linear model adjusted for age, sex, history of CKD, and maximum oxygen requirement at the time of blood draw using the Limma R package[53]. Protein expression data was log2 transformed.

For Supplementary Table 4 please see attachment.

**Supplementary Figure 1**

Protein quantitative trait loci (pQTL) evidence for 62 AKI-associated proteins. Associations were taken from proteomic and genomic data measured using human plasma samples.

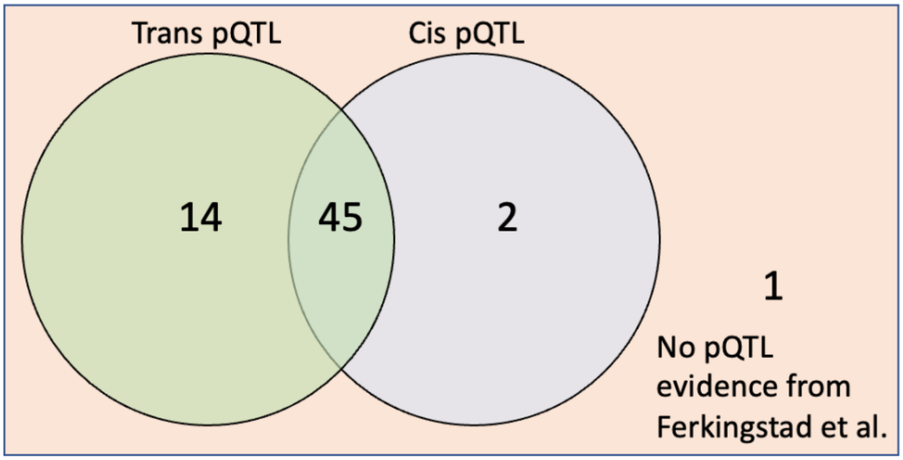


**Supplementary Figure 2**

Distribution of number of days after discharge for the first and last post-discharge eGFR measurements. All measurements were extracted from the EHR as part of routine clinical care.

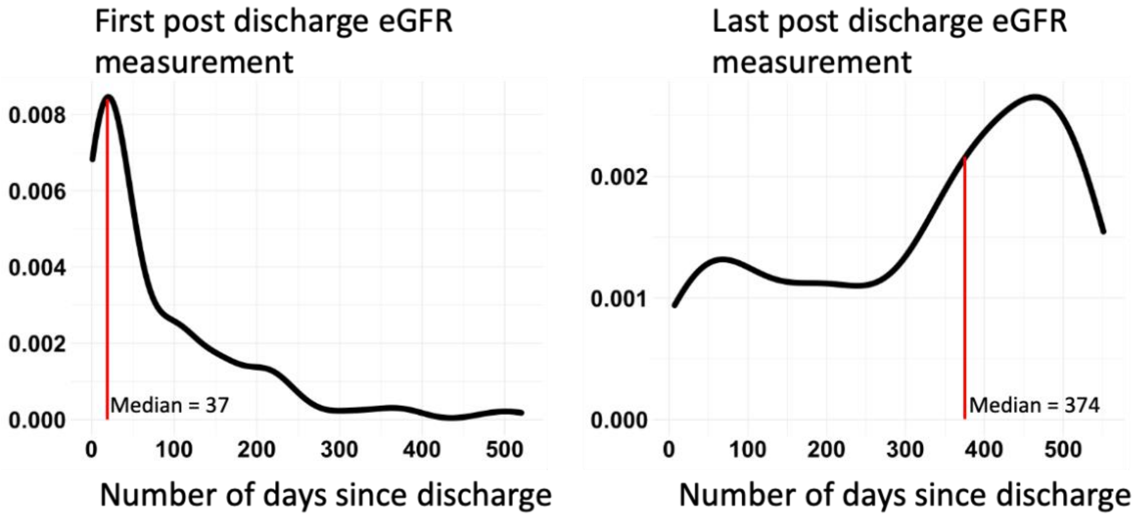
